# A Peptide-based Magnetic Chemiluminescence Enzyme Immunoassay for Serological Diagnosis of Corona Virus Disease 2019 (COVID-19)

**DOI:** 10.1101/2020.02.22.20026617

**Authors:** Xue-fei Cai, Juan Chen, Jie-li Hu, Quan-xin Long, Hai-jun Deng, Kai Fan, Pu Liao, Bei-zhong Liu, Gui-cheng Wu, Yao-kai Chen, Zhi-jie Li, Kun Wang, Xiao-li Zhang, Wen-guang Tian, Jiang-lin Xiang, Hong-xin Du, Jing Wang, Yuan Hu, Ni Tang, Yong Lin, Ji-hua Ren, Lu-yi Huang, Jie Wei, Chun-yang Gan, Yan-meng Chen, Qing-zhu Gao, A-mei Chen, Chang-long He, Dao-Xin Wang, Peng Hu, Fa-Chun Zhou, Ai-long Huang, Ping Liu, De-qiang Wang

## Abstract

A respiratory illness has been spreading rapidly in China, since its outbreak in Wuhan city, Hubei province in December 2019. The illness was caused by a novel coronavirus, named severe acute respiratory syndrome coronavirus 2 (SARS-CoV-2). Clinical manifestations related to SARS-CoV-2 infection ranged from no symptom to fatal pneumonia. World Health Organization (WHO) named the diseases associated with SARS-CoV-2 infection as COVID-19. Real time RT-PCR is the only laboratory test available till now to confirm the infection. However, the accuracy of real time RT-PCR depends on many factors, including sampling location and of methods, quality of RNA extraction and training of operators etc.. Variations in these factors might significantly lower the sensitivity of the detection. We developed a peptide-based luminescent immunoassay to detect IgG and IgM. Cut-off value of this assay was determined by the detection of 200 healthy sera and 167 sera from patients infected with other pathogens than SARS-CoV-2. To evaluate the performance of this assay, we detected IgG and IgM in the 276 sera from confirmed patients. The positive rate of IgG and IgM were 71.4% (197/276) and 57.2% (158/276) respectively. By combining with real time RT-PCR detection, this assay might help to enhance the accuracy of diagnosis of SARS-CoV-2 infection.

## Introduction

In December 2019, a novel coronavirus, labeled as severe acute respiratory syndrome coronavirus 2 (SARS-CoV-2) the Coronavirus Study Group [1], has been identified as the causative agent of the Corona Virus Disease 2019 (COVID-19) [2] outbreak in Wuhan, Hubei province of China [3, 4, 5, 6, 7]. This disease spread rapidly by human-to-human transmission [4] from Wuhan to other regions [8], resulting in more than 44,000 COVID-19 cases in mainland China based on the statistical data until Feb 12, 2020 [9]. Additionally, a total of 78 cases were identified in Hong Kong, Macao and Taiwan, and more than 400 cases were identified in Thailand, Japan, South Korea, United States, Vietnam, Singapore, Nepal, France, Australian and Canada [9]. So far, the number of infected people is still growing significantly.

Given that effective anti-viral therapeutics are unavailable currently, the first line of defense is to identify infected-patients as early as possible. Currently, laboratory diagnosis of SARS-CoV-2 has been carried out by detecting viral RNA in throat swab samples based on real-time reverse transcription polymerase chain reaction assay (real-time RT-PCR assay). This real-time RT-PCR method is sensitive and does not require live virus present in the specimen. Despite these advantages, such method can give results that are falsely-negative or falsely-positive due to several limitations, such as quality of specimen collection, multi-steps of RNA preparation and virus mutation. Another most widely used method is a serological test for the presence of antibodies against viral proteins. Like sever acute respiratory syndrome CoV (SARS-CoV) and Middle East respiratory syndrome CoV (MERS-CoV), SARS-CoV-2 are enveloped positive-sense single-stranded RNA viruses [5, 10]. Genomic analysis of SARS-CoV-2 reveals four major structural proteins including Spike (S) protein, Nucleocapsid (N) protein, Envelope (E) protein, and Membrane (M) protein, as well as a number of accessory open reading frame (ORF) proteins [5, 10]. In this study, we developed a magnetic chemiluminescence enzyme immunoassay (MCLIA) which has good specificity for immunological reaction and high sensitivity for detecting serum immunoglobulin G (IgG) and IgM against SARS-CoV-2.

## Materials and methods

### Human sera

A total of 276 sera were collected from 276 inpatients from three designated hospitals, Chongqing Three Gorges Central Hospital, Yongchuan Hospital Affiliated to Chongqing Medical University (CQMU), and The Public Health Center, in Chongqing, China. These patients were confirmed to be infected with 2019-nCov by real time RT-PCR detection of virus RNA. Among these sera samples, 168 were taken from patients with fever symptom. The time points of sampling range from day 2 to day 27 from the onset of fever. Ninety-nine of these patients reported exposure to persons with confirmed infection latter. The 200 normal human sera were collected from healthy people more than 1 year before 2019-nCov outbreak. 167 sera from patients with infection with other pathogens were collected from the Second Hospital Affiliated to CQMU and Children’s Hospital Affiliated to CQMU. These pathogens include influenza A virus (25), respiratory syncytial virus (7), parainfluenza virus (8), influenza B virus (5), adenovirus (6), *Klebsiella pneumonia* (8), *Streptococcus pneumonia* (3), *Mycoplasma* (5), *Acinetobacter baumannii* (10), *Candida albicans* (2), *Staphylococcus aureus* (3), *Mycobacterium tuberculosis*(4), Hepatitis B virus (33), Hepatitis C virus (22), Syphilis (23) and Saccharomycopsis (3).

All sera samples were inactivated at 56 °C for 30 min. This study was approved by the Ethics Commission of Chongqing Medical University (CQMU-2020-01). Written informed consent was waived by the Ethics Commission of the designated hospital for emerging infectious diseases.

### Synthetic peptide-based luminescent immunoassay

We developed a luminescent immunoassay for the detection of 2019-nCov antibody using synthetic peptide antigens as the immunosorbent. Twenty peptides, deduced from the genomic sequence from GenBank (NC_045512.1), were synthesized as candidate antigens from the orf1a/b, spike (S), and nucleocapsid (N) proteins. Each kind of peptide was labelled with biotin and the biotinylated peptide was purified and used to bind to streptavidin-coated magnetic beads. For antibody assay, serum samples (100μl/each sample) were mixed with the beads carrying corresponding peptides for 10 min at 37°C. Beads were washed 5 times, reacted to the antibody conjugate, again washed 5 times, and reacted to substrate. Reactivity was determined by a luminescence reader (Peteck 96-I, Bioscience, China).

### Evaluation of the luminescent immunoassay

Cut-off value of the test was determined as the mean luminescence value of the 200 normal sera plus 5 folds of SD. Sera from 276 COVID-19 patients 167 patients with irrelevant pathogens were used to evaluate the performance of the assay. The luminescent immunoassay was performed as described above. Results were determined as positive if the signal/cutoff (S/C) ratio ≥1.

## Result

### Evaluation of Synthetic peptide-based MCLIA for SARS-CoV-2

Twenty synthetic peptides, derived from the amino acid sequence of ORF1a/b, Spike (S) protein and Nucleocapsid (N) protein, were used to develop MCLIA for detecting IgG and IgM antibodies against SARS-CoV-2. To screen these peptides, 5 sera from confirmed patients and 10 normal sera were used to react with these peptides respectively. Among these peptides tested, one from S protein showed the best performance. We used the assay based on this peptide for the following study. To determine the cut-off value of this assay, serum samples from 200 healthy blood donors who donated blood 1-2 years ago were first tested. The mean chemiluminescence (CL) values for IgG and IgM were 0.152±0.109 and 0.151±0.107, respectively (Fig.1). Cut-off value for IgG and IgM detection were determined as 0.7 and 0.7 respectively.

**Fig 1.**
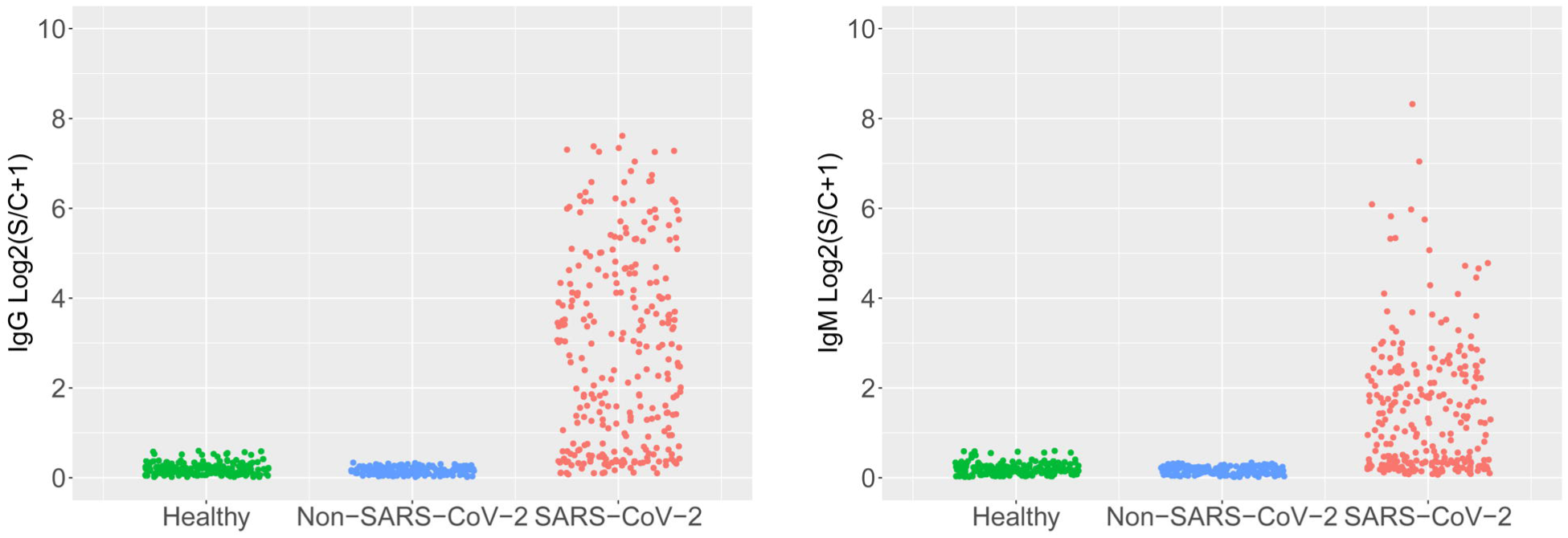
Evaluation of the synthetic peptide-based MCLIA for the detection of IgG and IgM against SARS-CoV-2. Serum samples were obtained from 200 healthy blood donors, 167 people infected with other respiratory pathogens and 276 patients with confirmed SARS-CoV-2 infection. The serum IgG (A) and IgM (B) were analyzed by MCLIA.

To test the specificity of the assays, the serum samples from 167 people infected with other respiratory pathogens such as influenza A virus, influenza B virus, parainfluenza virus, adenovirus, respiratory syncytial virus, mycoplasma, *Streptococcus pneumonia, Klebsiella pneumonia, Acinetobacter baumannii, Candida albicans, Staphylococcus aureus* were tested. The mean chemiluminescence (CL) values for IgG and IgM in non-SARS-CoV-2 infected people were 0.121 ± 0.062 and 0.120 ± 0.065, respectively (Fig.1A-B). These results showed that no cross-reactivity was observed for these 20 pathogens, indicating a very good specificity.

To test stability of this MCLIA based serological diagnosis method, serum samples with different concentrations were measured 10 times (Fig 2. A, B, C, D), coefficient of variation (CV) of IgG and IgM detection in different concentration samples were all below 6% (Fig 2. E, F), which meant a perfect stability of this assay in IgG/IgM detection. Furthermore, series dilutions for 6 serum samples(3 for IgG, 3 for IgM) were performed and S/co values were collected, regression analysis revealed S/co value range from 1 to 200 linear reflected serum antibody concentration (IgG, R^2^= −0.902, P<0.001, Fig 3A; IgM, R^2^= −0.946, P<0.001, Fig 3B), assured the rationality in further quantitative comparison based on S/co values.

**Fig 2.**
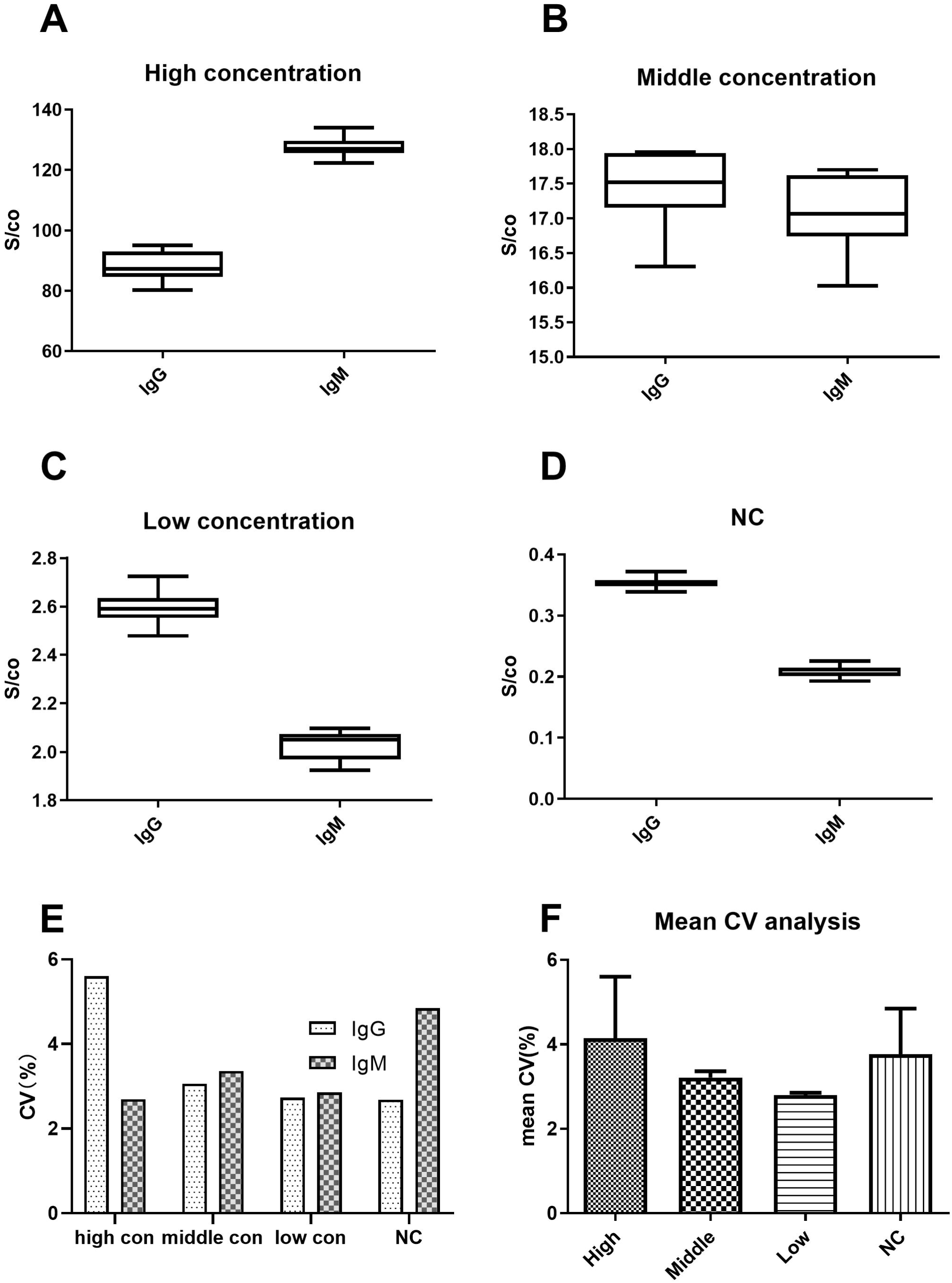
Assessment methodological quality of MCLIA. A. repeated detection analysis of high antibody concentration serum sample (time of replication=10). B. Detection analysis of middle antibody concentration serum sample (time of replication=10). C. Detection analysis of low antibody concentration serum sample (time of replication=10). D. Detection analysis of negative control(healthy) serum sample (time of replication=10). E. Percent coefficient of variation for different concentration serum samples. F. Mean percent coefficient of variation for the different concentration serum samples.

**Fig 3.**
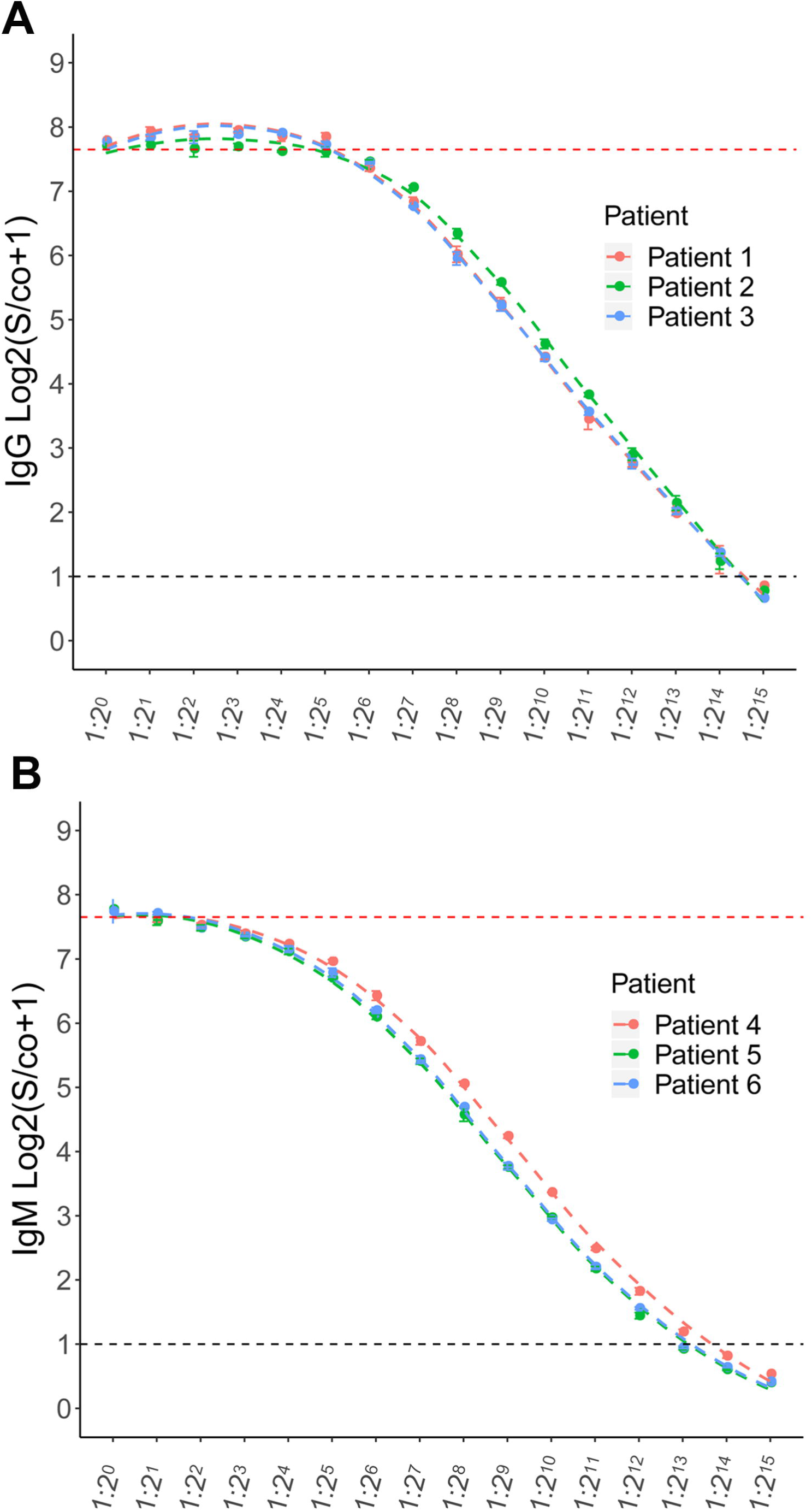
The correlation between the serial dilution ratio and calculated S/co values. A. correlation between serial dilution ratio and S/co values in IgG detection in 3 serum samples (*n*□=□3 replicates in each dilution for each sample). B. correlation between serial dilution ratio and S/co values in IgM detection in 3 serum samples (*n*□=□3 replicates in each dilution for each sample).

### Serological Diagnosis for SARS-CoV-2 in patients with confirmed infection

IgG and IgM were further examined by MCLIA in serum from 276 patients with confirmed SARS-CoV-2 infection. The median age of these patients was 48 years (IQR, 37-56; range, 0.66-84years), and 151/276 (54.71%) were men. The mean (CL) values for IgG were 18.62± 32.87, ranging from 0.05-194.56; IgM were 5.50±22.60, ranging from 0.04-318.16. Majority (197/276; 71.4%) of patients were positive for IgG antibody against the SARS-CoV-2, while 57.2% (158/276) patient were positive for IgM antibody against the SARS-CoV-2. Overall, 225 patients showed positive for IgM or IgG test and the total positive rate reached 81.52% (225/276) (Table 1).

**Table 1.** Positive rates of serum IgG or IgM in 276 patients with confirmed SARS-CoV-2 infection SARS-CoV-2, severe acute respiratory syndrome coronavirus 2

|  | Positive No. | Negative No. | Positive rate (%) |
| --- | --- | --- | --- |
| IgG | 197 | 79 | 71.37 |
| IgM | 158 | 118 | 57.24 |
| IgG or IgM | 225 | 51 | 81.52 |

## Discussion

We developed a luminescent immunoassay for IgG and IgM against SARS-CoV-2, which, to our knowledge, was the first such assay allowed us to study the antibody response to the newly identified coronavirus. This assay was based on a peptide from S protein, screened out from 20 candidate peptides deduced from the genomic sequence. Using synthetic peptide as antigen helps to enhance the stability and repeatability of an assay, and theoretically would be more specific than using virus as antigen. Indeed, this peptide showed a very good specificity in our assay: none of the 167 sera from patients infected with other pathogens than SARS-CoV-2 reacted with this peptide. This high specificity may be attributed to the relatively low homology of this region to other coronaviruses (data not shown).

Real time RT-PCR was the only test to confirm the infection of SARS-CoV-2 till now. We detected both IgM and IgG in the same sera from the 276 infection-confirmed patients. IgG was detected in 71.4% (197/276) of all the sera, higher than the detection rate of IgM (57.2%, 158/276). A combination the two antibodies enhanced the detection rate to 81.5% (225/276). Different sensitivity of the detection of IgG and IgM had been reported in SARS [11]. IgG can be detected as early as 2 days after the onset of fever. IgM was not detected earlier than IgG, similar to the situation in MERS [12], which limits its diagnostic utility. It was reported that 20%-50% SARS patients cannot be confirmed by RT-PCR [13], and this elicited a speculation that there might be a comparable part of infection cannot be detected by real time RT-PCR. Failure of detection by real time RT-PCR can be caused by the problems from sampling, RNA extraction and PCR amplification, while detecting antibodies in serum sample avoid a large part of these problems.

## Data Availability

All relevant data are within the paper

## Reference

1. Alexander E. Gorbalenya SCB, Ralph S. Baric, et al. Severe acute respiratory syndrome-related coronavirus: The species and its viruses – a statement of the Coronavirus Study Group. medRxiv. 2020. doi: https://doi.org/10.1101/2020.02.07.937862.

2. WHO. 2020. Available from: https://www.who.int/dg/speeches/detail/who-director-general-s-remarks-at-the-media-briefing-on-2019-ncov-on-11-february-2020

3. Chan JF, Yuan S, Kok KH, et al. A familial cluster of pneumonia associated with the 2019 novel coronavirus indicating person-to-person transmission: a study of a family cluster. Lancet. 2020 Jan 24. doi: 10.1016/S0140-6736(20)30154-9. PubMed PMID: 31986261.

4. Li Q, Guan X, Wu P, et al. Early Transmission Dynamics in Wuhan, China, of Novel Coronavirus-Infected Pneumonia. N Engl J Med. 2020 Jan 29. doi: 10.1056/NEJMoa2001316. PubMed PMID: 31995857.

5. Zhou P, Yang XL, Wang XG, et al. A pneumonia outbreak associated with a new coronavirus of probable bat origin. Nature. 2020 Feb 3. doi: 10.1038/s41586-020-2012-7. PubMed PMID: 32015507.

6. Huang C, Wang Y, Li X, et al. Clinical features of patients infected with 2019 novel coronavirus in Wuhan, China. Lancet. 2020 Jan 24. doi: 10.1016/S0140-6736(20)30183-5. PubMed PMID: 31986264.

7. Wei-jie Guan Z-yN, Yu Hu, et. al. Clinical characteristics of 2019 novel coronavirus infection in China. medRxiv. 2020. doi: http://dx.doi.org/10.1101/2020.02.06.20020974.

8. Yang Yang Q-BL, Ming-Jin Liu, et al. Epidemiological and clinical features of the 2019 novel coronavirus outbreak in China. medRxiv. 2020. doi: https://doi.org/10.1101/2020.02.10.20021675.

9. WHO. Coronavirus disease 2019 (COVID-19) Situation Report – 23. Available from: https://www.who.int/docs/default-source/coronaviruse/situation-reports/20200212-sitrep-23-ncov.pdf?sfvrsn=41e9fb78_2

10. Chan JF, Kok KH, Zhu Z, et al. Genomic characterization of the 2019 novel human-pathogenic coronavirus isolated from a patient with atypical pneumonia after visiting Wuhan. Emerg Microbes Infect. 2020 Dec;9(1):221–236. doi: 10.1080/22221751.2020.1719902. PubMed PMID: 31987001.

11. Woo PC, Lau SK, Wong BH, et al. Differential sensitivities of severe acute respiratory syndrome (SARS) coronavirus spike polypeptide enzyme-linked immunosorbent assay (ELISA) and SARS coronavirus nucleocapsid protein ELISA for serodiagnosis of SARS coronavirus pneumonia. J Clin Microbiol. 2005 Jul;43(7):3054–8. doi: 10.1128/JCM.43.7.3054-3058.2005. PubMed PMID: 16000415; PubMed Central PMCID: PMCPMC1169156.

12. Corman VM, Albarrak AM, Omrani AS, et al. Viral Shedding and Antibody Response in 37 Patients With Middle East Respiratory Syndrome Coronavirus Infection. Clin Infect Dis. 2016 Feb 15;62(4):477–483. doi: 10.1093/cid/civ951. PubMed PMID: 26565003.

13. Yam WC, Chan KH, Poon LL, et al. Evaluation of reverse transcription-PCR assays for rapid diagnosis of severe acute respiratory syndrome associated with a novel coronavirus. J Clin Microbiol. 2003 Oct;41(10):4521–4. doi: 10.1128/jcm.41.10.4521-4524.2003. PubMed PMID: 14532176; PubMed Central PMCID: PMCPMC254368.

